# From Variability to Standardization: The Impact of Breast Density on Background Parenchymal Enhancement in Contrast-Enhanced Mammography and the Need for a Structured Reporting System

**DOI:** 10.1101/2025.04.15.25325851

**Authors:** G Di Grezia, A Nazzaro, E Cisternino, A Galiano, G. Gatta, V Cuccurullo, M Scaglione

**Affiliations:** Department of Radiology, Link Campus University, Rome, Italy; REPRISE - Register of Expert Peer Reviewers for Italian Scientific Evaluation; Department of Radiology, P.O. ‘A. Perrino ‘Hospital, Brindisi, Italy; Department of Radiology, University of Campania “Luigi Vanvitelli”, Naples, Italy; Department of Nuclear Medicine, University of Campania “Luigi Vanvitelli”, Naples, Italy; Department of Radiology, University of Sassari, Sassari, Italy

**Keywords:** Breast Density, Background Parenchymal Enhancement, Contrast-Enhanced Mammography, BI-RADS, Risk Stratification, Standardized Lexicon, BPE-CEM Standard Scale (BCSS), Relational Data Base Management System (RDBMS), Applied Regression Analysis

## Abstract

**Introduction:** Breast density is a well-established predictor of breast cancer risk, affecting both the probability of malignancy and the sensitivity of mammography. Background parenchymal enhancement (BPE), which is observed in contrast-enhanced breast imaging, has been investigated as a potential independent biomarker for breast cancer risk. However, the literature presents conflicting results regarding its relationship with both breast density and malignancy. Unlike breast density, which is classified using the ACR BI-RADS system, BPE lacks a standardized reporting system in contrast-enhanced mammography (CEM), leading to inconsistencies in its assessment and clinical application. This study aims to determine whether breast density influences BPE in CEM and to establish a structured lexicon for BPE classification, which will enhance consistency in clinical practice and improve comparability across research studies.

**Materials and Methods:** This retrospective study included 213 patients who underwent CEM, mammography (MG), and ultrasound (US) between May 2022 and June 2023 at the P.O. ‘A. Perrino’ Hospital, Brindisi. BPE was categorized into four levels (Minimal, Light, Moderate, and Marked), while breast density was rated according to the ACR BI-RADS (A–D) classification. Statistical analysis was performed to assess the correlation between BPE and breast density, with the findings supporting the need for a BPE-CEM Standard Scale (BCSS) to standardize reporting and improve clinical consistency.

**Results:** Among the 213 patients, 57% exhibited minimal BPE, 31% light BPE, 10% moderate BPE, and 2% marked BPE. Higher breast density (ACR C–D) was significantly associated with increased BPE levels, while lower breast density (ACR A–B) correlated with lower BPE levels. Regression analysis confirmed a significant positive correlation (p < 0.05) between breast density and BPE, with no significant association found between BPE and age. These findings underline the necessity for a standardized BPE classification system, leading to the development of the BCSS, a structured lexicon aimed at enhancing clinical reporting and interobserver agreement.

**Conclusion:** BPE in CEM is significantly influenced by breast density, highlighting the need for a standardized classification system. To address the current variability in reporting, we propose the BPE-CEM Standard Scale (BCSS), which aims to ensure greater consistency in both clinical practice and research. Standardizing BPE characterization will facilitate its integration into breast cancer risk assessment models. Future research should focus on validating the BCSS framework and evaluating its impact on diagnostic accuracy and clinical decision-making.

## INTRODUCTION

Breast density is widely acknowledged as a crucial factor in predicting breast cancer risk. Higher breast density is associated with an increased likelihood of malignancy and a decreased sensitivity of conventional mammography [1,2]. Background parenchymal enhancement (BPE), a phenomenon observed in contrast-enhanced breast imaging, refers to the physiological uptake of contrast in non-pathological breast tissue. Extensive studies on BPE have primarily focused on breast MRI, where some research suggests that it may serve as an independent imaging biomarker for breast cancer risk, while other studies have found no significant correlation [3,4]. This lack of consensus hinders its widespread application in clinical practice.

Contrast-enhanced mammography (CEM) has emerged as a promising alternative to breast MRI, offering both anatomical and functional imaging, with advantages such as increased accessibility and shorter acquisition times [5,6]. Like MRI, BPE is also visible on CEM, although its expression differs due to the two-dimensional nature of mammographic imaging. In cases of pronounced BPE, lesion detectability may be compromised, potentially affecting diagnostic accuracy [7,8]. Despite its potential clinical relevance, the characterization and interpretation of BPE in CEM remain inconsistent, as no standardized classification currently exists. Unlike breast density, which is systematically classified using the American College of Radiology (ACR) BI-RADS system, the absence of a universally accepted lexicon for BPE in CEM results in variability in reporting and analysis across clinical and research settings [9].

The literature presents conflicting results regarding the relationship between BPE and breast density, as well as the correlation between BPE and breast cancer risk [10,11]. Some studies suggest that breast density may influence BPE, while others have found no significant correlation [12,13]. This inconsistency highlights the urgent need for a standardized and reproducible classification system for BPE in CEM, similar to the BI-RADS system used for breast density. Establishing a harmonized terminology would support more consistent data collection, improve interobserver agreement, and facilitate large-scale studies to evaluate the clinical significance of BPE [14,15].

### Study Objective

The aim of this study is to examine the relationship between breast density and BPE in CEM by analyzing a dedicated patient dataset. Additionally, we propose the BPE-CEM Standard Scale (BCSS), a structured lexicon designed to classify BPE in CEM. This four-tiered classification system—Minimal, Light, Moderate, and Marked— provides a structured and reproducible method for reporting BPE in both clinical practice and research. Establishing a standardized framework for BPE assessment will:

- Enhance interobserver agreement in CEM interpretation
- Facilitate more reliable comparisons across future studies
- Contribute to the standardized integration of BPE into structured reporting and risk assessment strategies

Through the development of a unified BPE lexicon, this study aims to improve the consistency and applicability of BPE evaluation in CEM, ultimately supporting more accurate breast cancer risk stratification and informed clinical decision-making.

## MATERIALS AND METHODS

### Study Design and Patient Selection

This was a single-center study conducted at the Interventional Senology Unit (UOSD) of P.O. ‘A. Perrino ‘Hospital in Brindisi between May 2022 and June 2023. The study adhered to Good Clinical Practice (GCP) guidelines. Since this was a retrospective study, informed consent was not required, except for the standard consent provided for imaging procedures.

A total of 314 patients were initially considered. After applying the exclusion criteria, 118 patients were excluded from the study. As a result, 213 patients aged between 28 and 80 years were included in the final analysis. These patients underwent ultrasound (US), mammography (MG), and contrast-enhanced mammography (CEM) within the study period.

The study included patients presenting with a suspicious breast lesion who underwent CEM prior to biopsy and subsequent surgery. Only patients with a histologically confirmed diagnosis of invasive breast cancer were included.

### Data Collection and Database Management

The data were collected and structured into three spreadsheets:

- Spreadsheet 1: Included patient names and dates of birth.
- Spreadsheet 2: Contained breast density classification (ACR A-D), Background Parenchymal Enhancement (BPE) grades, and three YES/NO columns indicating whether US, MG, and CEM were performed.
- Spreadsheet 3: Recorded millimeter-based measurements of the mammary glands in four numerical columns.

### Inclusion and Exclusion Criteria

#### Inclusion Criteria

- Age ≥18 years
- Positive CEM findings (BI-RADS 4-5 classification)

#### Exclusion Criteria

- Inability to undergo or complete CEM
- Poor image quality affecting diagnostic interpretation
- Absence of surgical specimen for histopathological confirmation
- Breast implants
- Previous diagnosis of breast cancer
- US- or stereotactic-guided biopsy performed within ≤21 days before CEM
- History of radiotherapy or systemic chemotherapy
- Contraindications to CEM, including:
  ∘ Pregnancy
  ∘ Hypersensitivity to contrast media
  ∘ Renal insufficiency, based on European Society of Urogenital Radiology (ESUR) guidelines

### Contrast-Enhanced Mammography (CEM) Protocol

All breast examinations were performed using Contrast-Enhanced Mammography (CEM) with standardized imaging protocols across all patients.

### Contrast Agent and Administration

Before intravenous (IV) administration of the contrast medium, patients were screened for potential risk factors associated with contrast reactions. Once confirmed that there were no contraindications, the contrast agent Iohexol 350 mg I/ml (Omnipaque®, GE Healthcare) was administered intravenously at a dose of 1.5 ml/kg through a 20G peripheral vein access, using a dual-syringe high-pressure injector at a flow rate of 3 ml/s. This was immediately followed by a 20 ml saline flush.

### Imaging Acquisition and Exposure Settings

- Premenopausal patients underwent the examination at any phase of the menstrual cycle without specific timing considerations.
- The mammary gland was compressed to acquire bilateral:
  ∘ Mediolateral oblique (MLO) views
  ∘ Craniocaudal (CC) views
- Imaging was performed approximately 2 minutes after contrast injection, and the procedure was repeated for the contralateral breast.
- Total imaging acquisition time was ≤7 minutes per patient.
- Low- and high-energy exposures were continuously acquired within 1.5 seconds under a single compression.

### Extended CEM Protocol

The CEM procedure was standardized using a full-field digital mammography system (Senographe Pristina with Seno Bright software, General Electric Healthcare®).

The imaging sequence included:

1. Compression of the healthy breast in the craniocaudal (CC) view, followed by acquisition of a low-energy (LE, 26–31 keV) and a high-energy (HE, 45–49 keV) image.
2. Compression of the affected breast in the CC view, with acquisition of LE and HE images.
3. Compression of the affected breast in the mediolateral oblique (MLO) view, with acquisition of LE and HE images.
4. Compression of the healthy breast in the MLO view, with acquisition of LE and HE images.
5. Compression of the affected breast in the mediolateral (ML) view, with acquisition of LE and HE images.
6. Compression of the healthy breast in the ML view, with acquisition of LE and HE images.

For each patient, a specific recombination algorithm was applied to subtract the LE and HE images, generating a single “subtracted” image for each projection.

No adverse effects were reported during the examinations.

Exposure parameters were adjusted based on breast size and glandular density, following a predefined value table. The images were analyzed using a high-resolution workstation (Barco, Belgium).

Low and high-energy exposures were continuously acquired within 1.5 seconds under a single compression. Each image was saved on the workstation in two formats:

- Low-energy images: Used to measure breast density.
- Recombined images: Used to assess BPE.

#### Image Interpretation

BPE was categorized into four levels: MIN (minimal), LIE (light), MOD (moderate), and MAR (marked).

Breast density was evaluated separately on the low-energy images according to the Breast Imaging-Reporting and Data System (BI-RADS) version 5 classification: A = predominantly fatty (<25% glandular); B = scattered fibroglandular densities (25–50% glandular); C = heterogeneously dense (51–75% glandular); D = extremely dense (>75% glandular).

All studies were interpreted by senior radiologists with expertise in breast imaging (at least 10 years of experience). BPE and breast density were paired for each patient and correlated with age and imaging results to identify potential trends.

### Statistical Analysis

The data were integrated into a relational database management system (DBMS) to reduce inconsistencies and redundancy [16, 17, 18].

The database comprised three tables reflecting the original spreadsheets, with primary and foreign key relationships established (Table 1, 2).

**Table 1.** Age group, record number, percentage, and density distribution by BPE.

|  |  |  |  |  |
| --- | --- | --- | --- | --- |
| Age Group | Number of Records (out of 268) | Percentage | BPE | Notes on Density |
| 25–40 | 11 | 5% | MIN/LIE |  |
| 25–40 | 2 | 1% | MOD/MAR | No A and no D |
| 41-55 | 80 | 38% | MIN/LIE |  |
| 41-55 | 13 | 6% | MOD/MAR | No A |
| Over 55 | 97 | 46% | MIN/LIE |  |
| Over 55 | 9 | 4% | MOD/MAR | No A, one B, and one D |

**Table 2.** Count of records by letter, with percentages and details on the S value.

|  |  |  |  |  |
| --- | --- | --- | --- | --- |
| letter | Count | Percentage (%) | Null Values | Details on S Value |
| A | 18 | 14% | 13 | 3 records with $S < -2$, 2 with $S > 2$ |
| B | 47 | 36% | 31 | 7 records with $S < -2$, 8 with $S > 2$ |
| C | 40 | 31% | 25 | 7 records with $S < -2$, 7 with $S > 2$ |
| D | 25 | 19% | 14 | 7 records with $S < -2$, 3 with $S > 2$ |

Queries were executed to perform analyses on:

- Patient age distributions based on birthdates and their associations with breast density and BPE.
- Grouping of BPE values into four categories (MIN, LIE, MOD, MAR).
- Distribution of breast density across 213 patients, including 313 imaging studies (both contrast-enhanced and non-contrast).
- Stratification of density and BPE by age groups (26–40, 41–55, over 55 years) using the MIN/LIE and MOD/MAR criteria.
- Comparison of lesion size (S) in millimeters between CEM and AP lesion measurements within the range of (-2,2) was conducted as suggested by Bland J.M., and Altman D.G., *Comparing methods of measurement: why plotting difference against standard method is misleading*, The Lancet, 346(8982), 1085-1087, 1995 [19].

A multiple linear regression analysis was conducted using Excel, with breast density as the dependent variable and BPE and age as independent variables. Numerical conversions of density and BPE were performed using conditional formulas to facilitate the analysis.

## RESULTS

1. Patient Age Distribution: Ages ranged from 28 to 79 years, with peaks among patients aged 60.
2. BPE Categorization: Among 211 patients, 57% exhibited MIN BPE, 31% LIE, 10% MOD, and 2% MAR.
3. Density and Imaging Modality: From 313 imaging studies:
  ∘ Without contrast: Density A (11%), B (29%), C (26%), D (17%).
  ∘ With contrast: Density A (1%), B (7%), C (4%), D (4%).
4. Age-Based Stratification: Density and BPE distribution by age group revealed:
  ∘ Ages 25–40: 5% MIN/LIE and 1% MOD/MAR.
  ∘ Ages 41–55: 38% MIN/LIE and 6% MOD/MAR.
  ∘ Over 55 years: 46% MIN/LIE and 4% MOD/MAR.
5. Difference Analysis (S): Of 213 records, 130 showed S values between -2 and 2, distributed as follows:
  ∘ Density A: 14% (3 S < -2; 2 S > 2).
  ∘ Density B: 36% (7 S < -2; 8 S > 2).
  ∘ Density C: 31% (7 S < -2; 7 S > 2).
  ∘ Density D: 19% (7 S < -2; 3 S > 2).

### Regression Analysis

The multiple linear regression yielded the following insights:

- Multiple R: 0.38, indicating moderate correlation.
- R^2^: 0.1443, showing that 14.4% of the variance in density is explained by BPE and age. In this case, only 14% of the variance in density is explained by BPE and age, which could correspond to a non-predictive model [20].
- Adjusted R^2^: 0.1361, suggesting limited impact from additional variables.
- Coefficients revealed a significant positive effect of BPE on density (p < 0.05) but no significant effect of age.
- Standard error is 0.8639. In this regard, it is important to consider, as suggested in Draper N.R., Smith H., *Applied Regression Analysis*, Wiley-Interscience, 2014 [21]; Altman D.G., *Practical Statistics for Medical Research*, Chapman & Hall, 1991 [22]; and Neter J., Wassermann W., Kutner M.H., *Applied Linear Statistical Models*, McGraw-Hill Education (ISE Editions), 1996 [23], that a lower standard error corresponds to greater accuracy in the model’s predictability (Table 3, 4).

**Table 3.** ANOVA – Analysis of Variance. This table presents the results of the ANOVA (Analysis of Variance) for the regression model. The table shows the degrees of freedom (df), sum of squares (SS), mean square (MS), F-statistic, and significance value for the regression and residual components, along with the total sum of squares. The very low significance F (7.86E-08) indicates that the model is statistically significant.

| ANOVA<br>(Analysis of Variance) |  |  |  |  |  |
| --- | --- | --- | --- | --- | --- |
| Source | df | SS | MS | F | Significance F |
| Regression | 2 | 26.42365998 | 13.21182999 | 17.70217459 | 7.8594E-08 |
| Residual | 210 | 156.7312696 | 0.746339379 |  |  |
| Total | 212 | 183.1549296 |  |  |  |

**Table 4.** Coefficients. This table displays the coefficients, standard errors, t-statistics, p-values, and 95% confidence intervals for the intercept and the predictor variables (BPE and Age) in the regression model. The coefficient for BPE is positive and statistically significant (p-value = 1.22E-07), while the coefficient for Age is negative but not statistically significant (p-value = 0.14).

|  | Coefficients | Standard Error | t Stat | p-value | Lower 95% | Upper 95% | Lower 95.0% | Upper 95.0% |
| --- | --- | --- | --- | --- | --- | --- | --- | --- |
| Intercept | 2.343692153 | 0.379884019 | 6.169493937 | 3.47353E-09 | 1.594817367 | 3.092566939 | 1.594817367 | 3.092566939 |
| BPEnum | 0.426517913 | 0.077858245 | 5.478134158 | 1.22301E-07 | 0.273034024 | 0.580001803 | 0.273034024 | 0.580001803 |
| Age | -0.008787179 | 0.005970236 | -1.471831024 | 0.14256352 | -0.020556454 | 0.002982096 | -0.020556454 | 0.002982096 |

### Regression Statistics

Multiple R: 0.379828178

R Squared: 0.144269445

Adjusted R Squared: 0.13612133

Standard Error: 0.863909358

Observations: 213

Overall, the regression analysis demonstrated that BPE has a significant positive effect on breast density, while age does not exert a statistically significant impact. Additionally, only a fraction of the variance in the dependent variable (breast density) is explained by the model, suggesting the potential influence of other variables not included in the analysis [21].

Based on the error standard value 0.8639 (Mean Squared Error: MSE, metric), it is reasonable to assume that a more predictive model could be implemented by studying the MAE (Mean Absolute Error) metric using AI techniques, as well-known deep learning provides more accurate predictions.

## DISCUSSION

The relationship between background parenchymal enhancement (BPE) and breast density has been the subject of considerable debate, with conflicting findings regarding their correlation and respective roles in breast cancer risk stratification [22, 23]. BPE, initially characterized in contrast-enhanced breast MRI, represents the physiological uptake of contrast in non-pathological breast tissue. This phenomenon is influenced by various factors, including hormonal status, vascularization, and glandular composition [24]. While some studies have proposed that BPE could serve as an independent imaging biomarker for breast cancer risk, others have reported inconclusive or contradictory findings [25]. Similarly, breast density is a well-established predictor of breast cancer risk, with higher-density breasts associated with an increased likelihood of malignancy and reduced sensitivity in conventional mammography [26, 27]. However, the interaction between BPE and breast density, particularly in contrast-enhanced mammography (CEM), remains unclear, largely due to the absence of a standardized system for assessing BPE in this imaging modality.

CEM, an advanced technique that combines anatomical and functional imaging, has gained recognition as a promising alternative to MRI, particularly for women with dense breasts, who may experience limited sensitivity on conventional mammography [28]. Like MRI, CEM visualizes BPE, but its expression differs due to the two-dimensional nature of mammographic imaging. This variability in the presentation of BPE across imaging modalities complicates both its interpretation and clinical utility [29]. Unlike breast density, which is systematically classified using the ACR BI-RADS system (A–D), no universally accepted terminology exists for BPE in CEM. This lack of standardization leads to inconsistencies in data interpretation, interobserver agreement, and comparative research [30].

Given these challenges, our study aimed to examine the relationship between breast density and BPE in CEM and to propose a structured classification system for BPE reporting. By analyzing real-world clinical data, we provide evidence supporting a significant correlation between higher breast density and elevated BPE levels in CEM, emphasizing the necessity for standardized reporting criteria.

### Key Findings and Clinical Implications

Our results confirm that patients with higher breast density (ACR C–D) exhibit increased levels of BPE, while those with lower breast density (ACR A–B) show lower BPE levels. Specifically:

- 57% of patients exhibited minimal BPE, 31% light BPE, 10% moderate BPE, and 2% marked BPE.
- A significant positive correlation was found between breast density and BPE, suggesting that fibroglandular tissue plays a crucial role in the manifestation of BPE.
- No statistically significant association was observed between BPE and patient age, reinforcing the hypothesis that breast density is a stronger determinant of BPE levels than age alone.
- Regression analysis further confirmed that breast density influences BPE, supporting the need to consider both parameters when interpreting CEM results.

These findings align with studies reporting a direct relationship between breast density and BPE [22], yet diverge from others that found no significant correlation [23]. This inconsistency highlights a broader issue: the lack of a standardized method for evaluating BPE in CEM.

### Need for a Standardized Lexicon for BPE in CEM

To address this issue, we propose the BPE-CEM Standard Scale (BCSS), a structured lexicon designed to offer a clear and reproducible method for BPE categorization in CEM reports, ensuring standardization across both studies and clinical practice. This scale would enhance consistency in reporting, facilitate interobserver agreement, and enable more reliable comparisons across future studies assessing BPE in CEM.

### Limitations and Future Directions

While our study provides valuable insights into the relationship between BPE and breast density, it is not without limitations, including the retrospective design, the mixed screening and diagnostic population, and the lack of data on menstrual cycle status [23]. Further research involving larger prospective cohorts is essential to validate our findings and refine the BCSS system for broader clinical implementation.

## CONCLUSION

Our findings highlight the urgent need for a standardized approach to BPE classification in CEM. Given the association between higher breast density and increased BPE, incorporating both parameters into breast cancer risk models could enhance personalized screening strategies.

The introduction of the BPE-CEM Standard Scale (BCSS) marks a significant step toward standardizing BPE reporting. This framework could lead to better risk stratification and improved diagnostic accuracy. Future prospective studies are required to validate this classification system and explore the potential of BPE as an independent biomarker for breast cancer risk.

## Data Availability

All data produced in the present study are available upon reasonable request to the authors

